# Allelic Diversity, de novo CAG Expansions, and Intergenerational Instability at the HTT Locus in a clinical sample of Huntington’s Disease from India

**DOI:** 10.1101/2025.03.03.21260193

**Authors:** Nikhil Ratna, Sowmya Devatha Venkatesh, Swathi Pasupulati, Geetanjali Murari, Nitish Kamble, Biju Viswanath, Ravi Yadav, Mathew Varghese, Pramod Kumar Pal, Sanjeev Jain, Meera Purushottam

**Affiliations:** Molecular Genetics laboratory and Genetic Counseling and Testing Clinic (GCAT), Department of Psychiatry, National Institute of Mental Health and Neurosciences, Bengaluru, 560029, India; Department of Neurology, National Institute of Mental Health and Neurosciences, Bengaluru, 560029, India

**Keywords:** Huntington’s Disease, CAG repeat, intermediate allele, reduced penetrance alleles, homozygous expansions, Juvenile HD, CAG instability, biallelic expansions, genetic modifiers

## Abstract

**BACKGROUND:** Huntington’s disease (HD) is an inherited, neurodegenerative disorder, caused by the expansion of an unstable CAG repeat sequence in the HTT gene. The prevalence of HD, allelic diversity, rate of novel expansions, and the clinical correlates, vary across populations.

**OBJECTIVE:** We aimed to analyze the diversity of alleles, and their clinical correlates; and describe the mode of inheritance and the pattern of instability of CAG repeats in a few families

**METHODS:** Clinical history and pedigree structure were collected from clinical records, or through interviews between 2016-19. Genetic testing at the HD locus was done on clinical suspicion, or relatedness, after counseling. Descriptive statistics and correlation analysis were used.

**RESULTS:** Expanded repeats were detected in 239 individuals, including 232 who were symptomatic, and seven presymptomatic relatives. The number of CAG repeats (mean=45.6) and age at onset (AAO) (mean=39.2 years) showed a strong inverse correlation (r=-0.67). We found atypical alleles such as 8 intermediate alleles (IA), 12 reduced penetrance alleles (RPA) and 14 large (>60) expansion alleles corresponding to juvenile HD. Three individuals carried biallelic expansions. Paternal inheritance was more common and the mean increase in repeats in the available parent-child pairs was 14. Thirty-seven individuals had no family history of HD, of which a *de novo* expansion could be ascertained in 3 cases.

**CONCLUSIONS:** Novel mutations at HTT locus may not be rare in India. A lack of family history should not exclude appropriate testing. Prevalence of intermediate alleles and incidence of de novo expansions, suggests that there may be a reservoir of alleles prone to expansion.

## Introduction

Huntington’s disease (HD) is a progressive neuro-degenerative disorder clinically characterized by a spectrum of motor, behavioral and cognitive symptoms. HD is caused by a triplet (CAG) repeat expansion in exon 1 of the *Huntingtin* (*HTT*) gene on chromosome 4 [OMIM 613004] and inherited in an autosomal dominant manner (1) . Alleles with CAG expansions >39 are fully penetrant (FPA), while alleles with 36-39 repeats are called reduced penetrance alleles (RPA) as they may or not develop any symptoms in their lifetime. Alleles with repeats between 27-35 called intermediate alleles (IA), do not cause HD but are highly unstable and prone to pathogenic expansions (>35) in the offspring (2). The length of the CAG repeat inversely correlates with age at onset (AAO) of motor symptoms (3). Other genetic and environmental modifiers can also modify the AAO and the outcome (4). The prevalence of HD, at-risk haplotypes, intermediate alleles, and the incidence of *de novo* mutations seem to vary across populations, suggesting possible differences in the extent of CAG instability (5–9). Previous studies have reported on CAG instability during transmission and the prevalence of latent risk of HD posed by IAs and RPAs (10,11). Despite reports suggesting that HD is not uncommon in India, there is sparse data on the borderline expansions, and their transmission trends across generations. We analyzed a well-characterized family-based cohort and report the demographics, inheritance patterns, instability trends and pertinent clinical correlates.

## Materials and Methods

A total of 390 screened individuals at the National Institute of Mental Health and Neurosciences (NIMHANS) between 2011-2019, underwent genetic testing for the HD mutation, and 239 were positive (Fig 1). Subjects without clinical data (12) and pre-manifest individuals (7) were excluded from analysis, leaving 220 individuals for the AAO based analysis (Fig.1). The study was approved by the Institute Ethics Committee (Ref No. NIMH/DO/ETHICS SUB-COMMITTEE 29th MEETING/2016). A written informed consent was obtained from all the study subjects, or their representatives. Both pre and post-test genetic counseling was offered to all individuals. Demographics, clinical details and pedigree charts were obtained from medical records and prospective clinical assessments. For pedigrees with no family history, more details were obtained through telephonic interviews. To understand the CAG-AAO trends within and across families, parent-child, sibling-sibling pairs were identified and analyzed separately. Sub-group analysis was also done for juvenile and sporadic cases. PCR specifications are provided in the supplementary file. STR analysis by Gene Print 10 system (Promega) was carried out to confirm 50% allele sharing in families with de novo expansions. Descriptive statistics, correlation analysis, and group comparisons were used as needed.

**Figure1:**
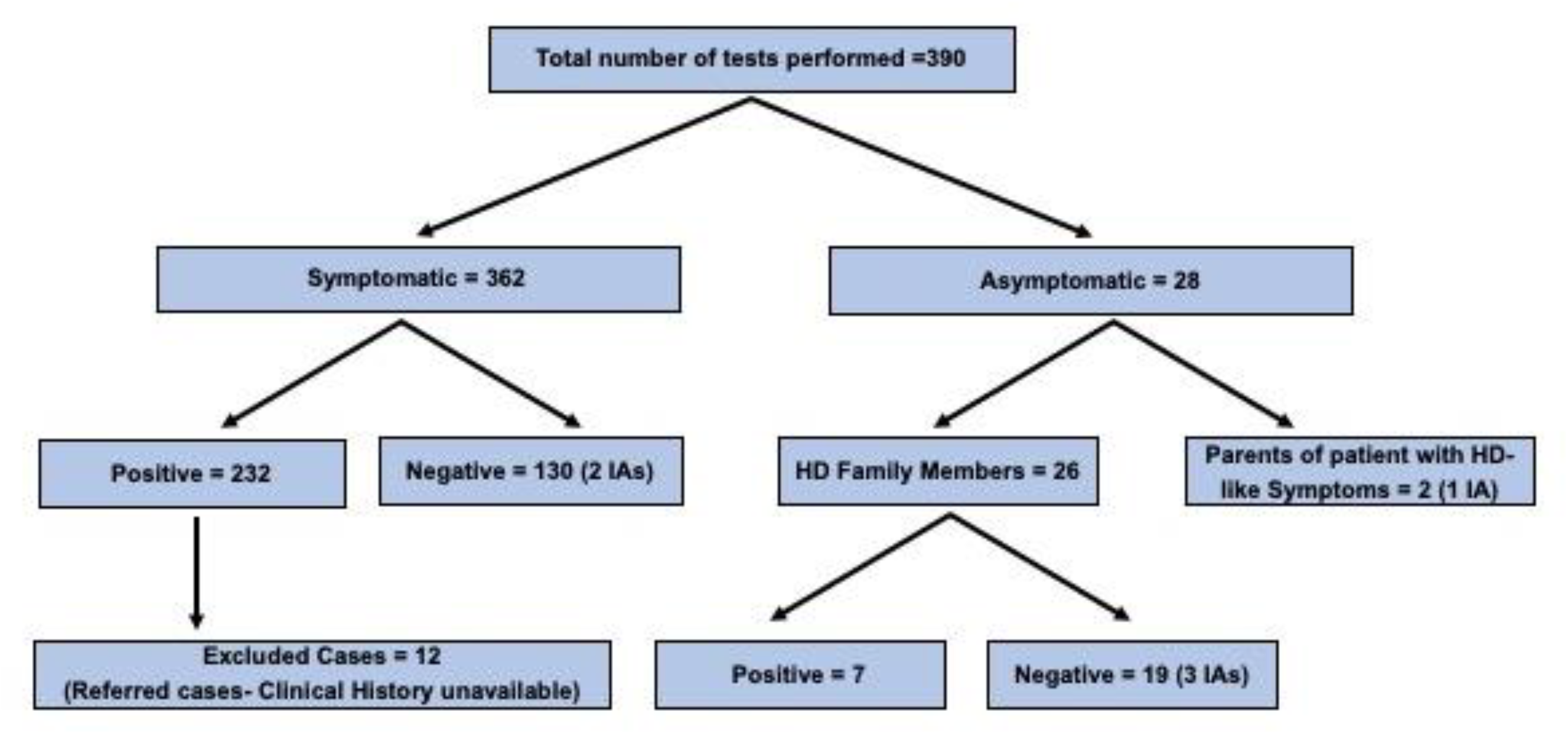
Summary of HD testing and results in the study duration. IA=Intermediate allele

## Results

### Demographics, AAO and CAG repeat length

The mean AAO of these subjects (N 220) was 39.2 ± 12.7 (**Range 4-72**) years. Most individuals were from southern and eastern India, but several were from other parts of India (supplementary fig 1). The genders were almost equally represented, with a slight male preponderance (F 44%; M 56%). Mean upper and lower allele repeat lengths were 45.6 ± 8.3 repeats (range 38-113) and 17.9 ± 2.8 repeats (range 9-37) respectively. (Table S1, S2). The AAO showed a strong negative correlation with the longer allele (r = - 0.67, p < 0.001) (Figure 2B). There were 26 families with more than 1 affected individual in this cohort.

**Figure 2:**
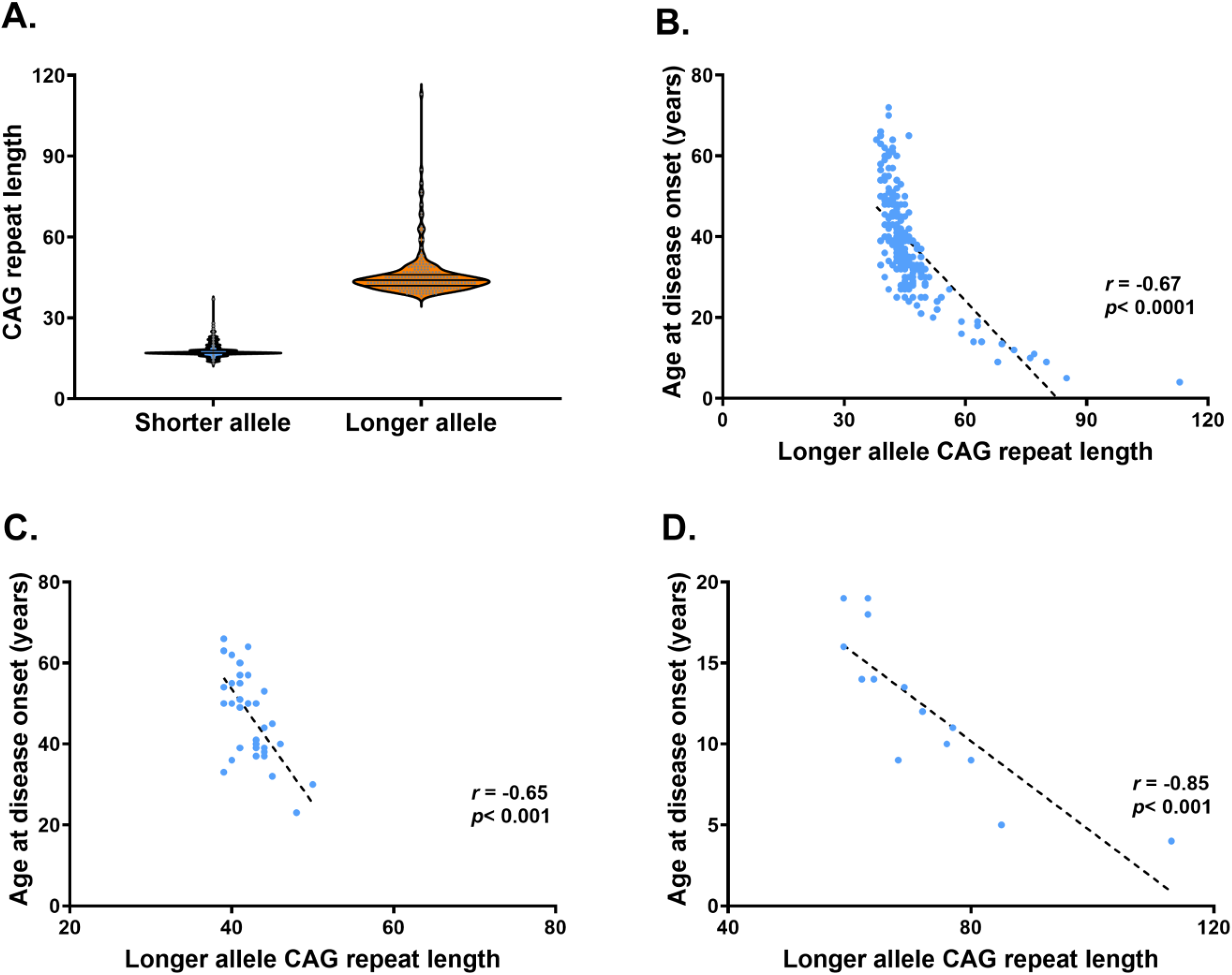
A) Distribution of longer and shorter alleles in HD patients (*n* = 220). (B–D) Correlation between CAG repeat length and Age at Onset (AAO) in: .(B) The entire cohort .(C) Family History Negative (FHN) cases .(D) Juvenile HD cases

### Reduced penetrance alleles (RPA), Intermediate alleles (IA), and Biallelic expansions (BE)

In this cohort, we found 12 patients with RPAs, and seven of these had behavioral onset with a psychiatric syndrome as the primary diagnosis. Half of these cases (6/12) did not have a family history of chorea. We also found 8 individuals with IAs (27-35) in our cohort. In two instances, an intermediate allele accompanied a fully penetrant allele. In another three instances, the IAs were detected as longer alleles in the asymptomatic parents or siblings of HD patients with a de novo expansion (supplementary Table 6). In addition, in two young people (20-30yrs) we detected to have IAs, who showed HD-like symptoms, and in whom alternative diagnoses like Acanthocytosis, Wilson’s disease and metabolic causes had been ruled out. Both showed evidence of caudate atrophy on MRI. Interestingly, a healthy parent of one of them also had an IA, an unexpected finding.

We also detected 3 cases of biallelic-expansions (28/43, 27/47, 37/41), but without consanguineous parentage. In one biallelic individual (37/41), there was no history suggestive of HD either, as one parent died prematurely (4^th^ decade) of unknown cause, and the other parent was alive and healthy (in 6^th^ decade). These biallelic cases were clinically indistinguishable from the rest.

### Juvenile-Onset HD and large expansion alleles

Fourteen (8 F) individuals of the cohort had juvenile onset HD (JHD), having developed symptoms at quite a young age of 12±4.5 years. Mean CAG repeat length was 72.1±14.2. All of them, except one, were paternally transmitted. We found 2 JHD sibling pairs in this cohort. The highest CAG length observed was 113. The correlation between repeat number and AAO was more pronounced than in adult patients (r=-0.85, P<0.001) (Fig 2d).

### Inheritance pattern and family history negative (FHN) cases

Most patients (170 out of 220) had at least one parent affected with HD, with paternal inheritance (95, 55%) being slightly more common. The mean repeat length and dispersion of the expanded allele was higher in those with paternal transmission as compared to those with maternal transmission (48.1 ± 11.5 vs 44.7 ± 4.3, p=0.01). However, there was no difference in AAO between the groups (36.2 ± 14.6 vs 38.9 ± 9.9 years, p=0.19).

Of the remaining 50, family history was unavailable in 13 cases as they were orphaned or adopted at a young age. In the remaining 37 of them neither of the parents, nor parent’s siblings were known to have HD. We refer to them as family history negative (FHN) cases. In 26 FHN cases, both parents had lived at least 60 years of life with no symptoms; while in the remaining 11 FHN cases, one parent had died in middle-age, but had no symptoms to suggest HD, and the surviving parent (>60) too had no symptoms. However, in six instances, one parent had late-onset (6^th^-8th decade) behavioral changes, but no motor symptoms. There were two cases with HD symptoms in siblings, but both parents lived past 60 years without showing HD symptoms. (supplementary table 3). In FHN cases, the mean size of the expansion was lower (42 repeats), and the age at onset was higher (46 years), but the correlation between repeat number and AAO (r=0.66, p<0.001) was similar when compared to those who had an identifiable family history (Figure 2C).

We could test family members in 3 FHN cases where parents/sibling were available and consented for testing after the genetic counseling. In two of them, IAs (32 and 30 repeats) from asymptomatic parents, had progressed to de novo mutations (43 and 44 repeats) in the offspring. In the third case with 48 repeats, the sibling had 32 repeats, while the living parent had normal alleles. The other parent had died in 7^th^ decade, but never had any symptoms of HD. We suspect that this parent might have had an IA, which was stably transmitted in the case of sibling, but expanded in the patient. We also note that the sibling with IA had generalized anxiety. STR genotyping confirmed paternity and sibship in the three families described. Overall, a significant proportion (16%) of the cases in this cohort had apparently novel expansions.

### CAG repeat expansion trends within a few families

From 26 families, we could identify 23 sibling pairs and 13 parent-offspring pairs (excluding de novo expansion pairs) with expanded repeats, of which four individuals were pre-symptomatic. Among sibling pairs, there were 15 paternal, 7 maternal and 1 unknown transmission (foster parents): (supplementary table 4). Repeat length of the expanded alleles and the AAOs strongly correlated between affected siblings (r=0.95, 0.85 respectively), with a lesser mean AAO of 4.5 years in younger sibling than elder sibling. (Figure 3a and 3b).

**Figure 3:**
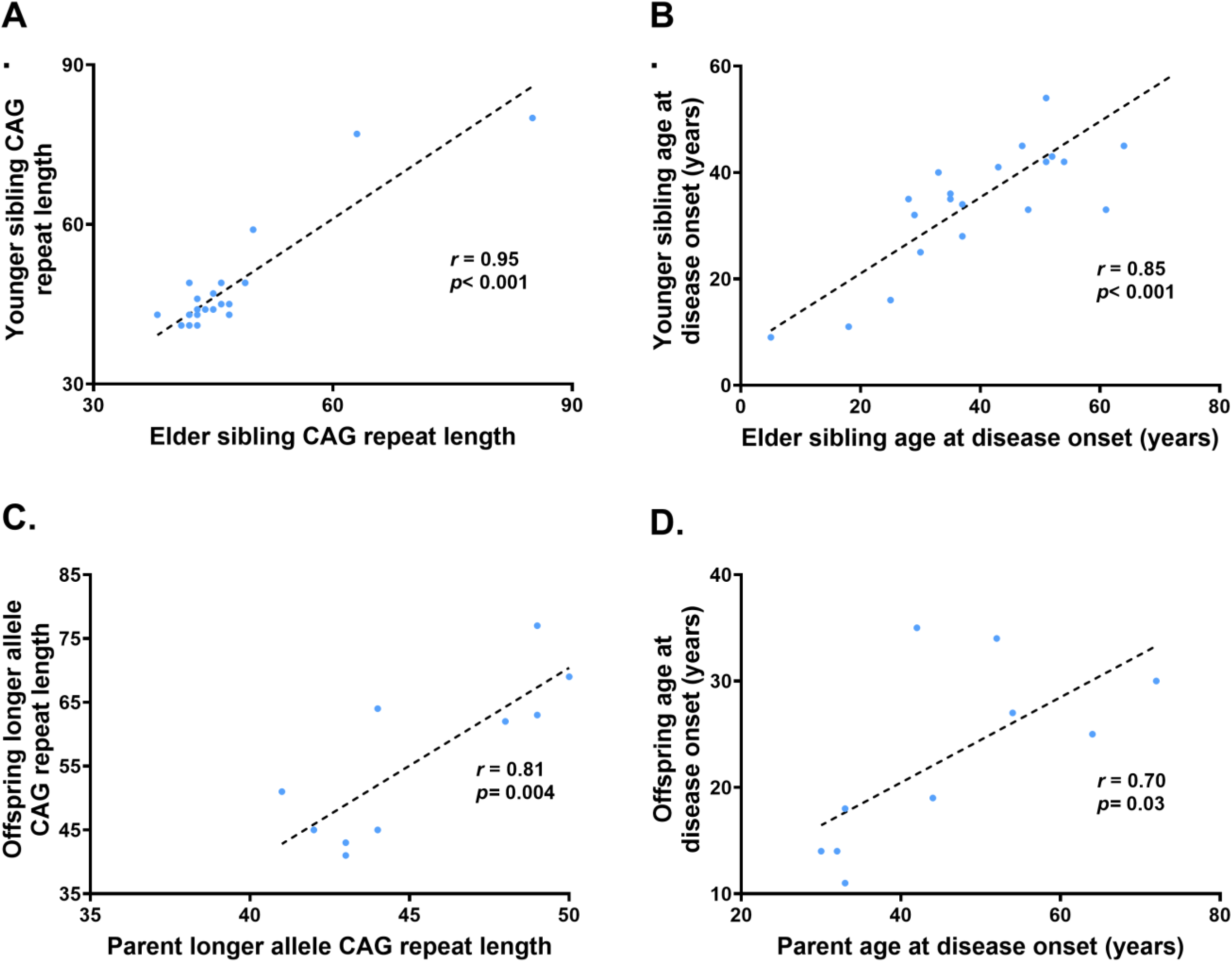
A. Correlation plot of CAGs of sibling pairs, B. Correlation plot of AAOs of sibling pairs, C. Correlation plot of CAGs of parent-offspring pairs, D. Correlation plot of AAOs of parent-offspring pairs

Among the 13 parent-offspring pairs (including 5 JHDs), the expanded allele had increased by about 14 repeats from parent (44.6 ± 3.1) to offspring (58.15 ± 20.4). Eight were paternal transmissions, while five were maternal. In 9 pairs, there was an expansion in repeats (Range 1-70); 2 pairs did not show any expansion, while 2 pairs showed contractions of one or two repeats from parent to offspring (supplementary table 5). Mean AAO of the parent and the offspring were 45.6 ± 14.6 (30 – 72) and 22.7 ± 9.2 (11 – 35) years respectively, with a mean difference of 23 years (7-42 years). CAG repeats and AAOs of parents correlated strongly with that of offspring (r=0.81, p=0.001; r=0.70, p=0.01) respectively (figures 3c and 3d).

## Discussion

Despite several studies of HD in India, the lack of data regarding transmission and distribution of expanded alleles within families, and the occurrence of de-novo expansion, in the population, was the motivation for this study. We found strong evidence for CAG instability in this small set of families, and the prevalence of a range of alleles from intermediate alleles to very large expansions. Incidentally we found intermediate alleles in young individuals suspected to have HD without any family history of HD. We present the insights derived from these findings to inform epidemiological research and clinical diagnosis.

### Cohort characteristics and trends in families

The overall distribution of ages-at-onset, and CAG repeat sizes, and their correlation was broadly identical with those reported earlier from India (12–14) and from elsewhere (3,15). The variations discovered in this well-documented cohort, and their familial and genetic characteristics, promotes a more nuanced understanding of HD in south Asia.

Interestingly, 2 individuals with atypical clinical syndromes and symptoms of chorea had repeat sizes in the IA range. Whether IAs may be clinically innocuous, or contribute to risk in some ways, is thus still a matter for investigation (16,17). Overall, out of 5 cases of IA transmission from 4 parents, 2 of them were stably transmitted, whereas 3 of them expanded to pathogenic range in the offspring. Variable instability of IAs has reported earlier(18). Somatic instability in symptomatic IA carriers revealed that IAs expand somatically in a CAG and age dependent manner, but whether this expansion, or other genetic predisposition lead to disease is still unclear (19).

Correlation of repeat sizes between siblings was stronger than in parent-offspring pairs. This suggests that the major expansion event perhaps occurs prior to gametogenesis (3), whereas marked variation within some sibling pairs could be due to downstream events. In two-thirds of sibling pairs, younger siblings had an earlier onset, suggesting that the parental age at transmission influences anticipation. Although anecdotal, marked variation of AAO in siblings with similar repeats suggests a significant role of disease modifying genetic variant in these cases. As expected, anticipation was high with paternal transmission, the most prominent one was seen in a JHD case (AAO-4 years, CAG-113) whose parent (CAG-43) was asymptomatic (20). Such extremely large expansions have been observed in JHD, as when a child had more than 200 repeats whereas the father had 43 (21). Whether these are a part of an inherent bias in chromatin organization or chromosomal arrangements, or other genetic and (or) environmental modifiers, needs to be explored.

### FHN cases, atypical alleles and de novo expansions-epidemiological implications

In almost a sixth of the patient sample, there was no obvious family history of an affected relative; but in a few cases, one of the parents developed late-onset behavioral symptoms, perhaps a manifestation of RPAs. Half (6/12) of the patients with RPAs had a spectrum of psychiatric symptoms, with mild or no motor symptoms. Such association of modestly expanded alleles with psychiatric symptoms has been reported (22). In addition, asymptomatic parents or siblings were detected to have an IA, while the index patient had an expanded allele, as has also been reported earlier (10,11). Such prevalence of IAs in the family members of FHN cases pose a risk for de novo expansions in their offspring. The prevalence of ‘sporadic’ HD differs across populations (23,24), owing to the varying incidence of de novo expansions of the IAs reported earlier (0%-14%) (25,26). The range of CAG repeats (39-50) and mean AAO of 46 in FHN cases of this sample suggests that clinicians should suspect HD in young individuals, even in the absence of family history.

The prevalence of IAs is reported to be about 6% in the general population, and in 7% of those with HD (27). The incidence of disease-causing expansions has been shown to correlate with the distribution of repeat sizes, the prevalence of IAs, and at-risk haplotypes, thus influencing the overall HD prevalence rates in European, American, Asian, and African populations (7). Exploring haplotypes of expanded alleles from FNH cases and in cases of other large expansion transmissions could shed light on the cis-acting factors contributing to expansion. In the 1000-genome data cohort, seven healthy (presumably) individuals who had pathogenic expansions (36-52 repeats) were detected in 2486 healthy individuals (28). A study from Ecuador reported a high prevalence of IAs (18%) with significant ethnic differences (29); while extremely short alleles as well as intermediate alleles were seen in a northern European non-HD population (30). The occurrence of JHD alleles (6%) in this cohort is comparable with cohorts from elsewhere, but much higher than other reports from India. A testing center from India reported the presence of 250 expanded alleles, including 9 RPAs, 7 large alleles >60 repeats, and 9 IAs (in negative samples) from 503 individuals who were tested, though clinical and family history details were not described (12). The epidemiology of intermediate and reduced penetrance alleles, and possible intersection with clinical symptoms, in the population thus needs investigation.

Bi-allelic expansions in three individuals in our sample could reflect the predisposition to allelic expansion in the general population, given the lack of consanguinity and at least one parent not having HD in all 3 cases. However, given the high rates of endogamy in southern India, the added risk for bi-allelic expansions and homozygosity in the subsequent generations of heterozygous patients’ needs to be noted. Significantly early age of marriage and reproduction (relative to age at onset) in lower socio-economic strata, further increases this risk.

### Limitations

Our study, based at a tertiary referral hospital, has an ascertainment bias, and may lack sufficient statistical power to generalize the findings. However, as HD is a rare disorder, our data highlight the importance of family-based analysis to understand the transmission dynamics, rate of de novo expansions and the prevalence of late onset psychiatric syndromes in such families.

## Conclusions

Our study adds to the fund of existing knowledge of HD genetic epidemiology in India and brings some novel perspectives. Our findings suggest that novel expansions may not be rare, and there could be a load of expansion prone alleles (IAs/RPAs) prevalent in Indian population. This has significant epidemiological implications. Hence, there is an urgent need for both diagnosis-driven and population-based prevalence studies to estimate the current and projected burden of HD in India. Family-based genomic studies would identify genetic modifiers of CAG instability, understand disease pathology and may even help develop personalized treatments.

## Data Availability

The data presented in the paper is available with Dr Nikhil Ratna

## Acknowledgments and Funding

HD patients were seen at the Movement Disorder Clinic and the Genetic Testing and Counseling Clinic at the National Institute of Mental Health and Neurosciences, Bengaluru, India. Material support for the work was provided by the Molecular Genetics Laboratory.

Dr Nikhil Ratna was supported by the Indian Council of Medical Research (ICMR) MD-PhD fellowship. Participants were included in the study after obtaining consent, and institutional ethics clearance. Ref No. NIMH/DO/ETHICS SUB-COMMITTEE 29th MEETING/201

