## Supplemental material for "Allelic Diversity, de novo CAG Expansions, and Intergenerational Instability at the HTT Locus in a clinical sample of Huntington’s Disease from India"

**Supplementary material**

**Methods-PCR specifications**

The genomic DNA was extracted from whole blood by salting out method. This DNA was used for polymerase chain reaction (PCR) to amplify the HTT locus using appropriate primers. DNA extracted from whole blood by salting out method was processed using polymerase chain reaction (PCR) with appropriate primers to amplify the *HTT* locus.

FP: 5’ -/5 HEX/ GGGATCGCGATGGCGACCCTGGAAAAGCTGATGAA 3’

RP: 5’ GGCGGTGGCGGCTGTTGCTGCTGCTGCTGC 3’

The occurrence of CAG expansions was identified by high resolution agarose gel electrophoresis (gel percentage is 2.5%) of the amplified products. For accurate estimation of amplicon size, fragment analysis was performed using ABI^TM^ 3500XL Genetic analyzer (Life Technologies^TM^, California, USA). Modal CAG repeat lengths for both alleles were determined using GeneMapper^TM^ v3.5.

**Results**

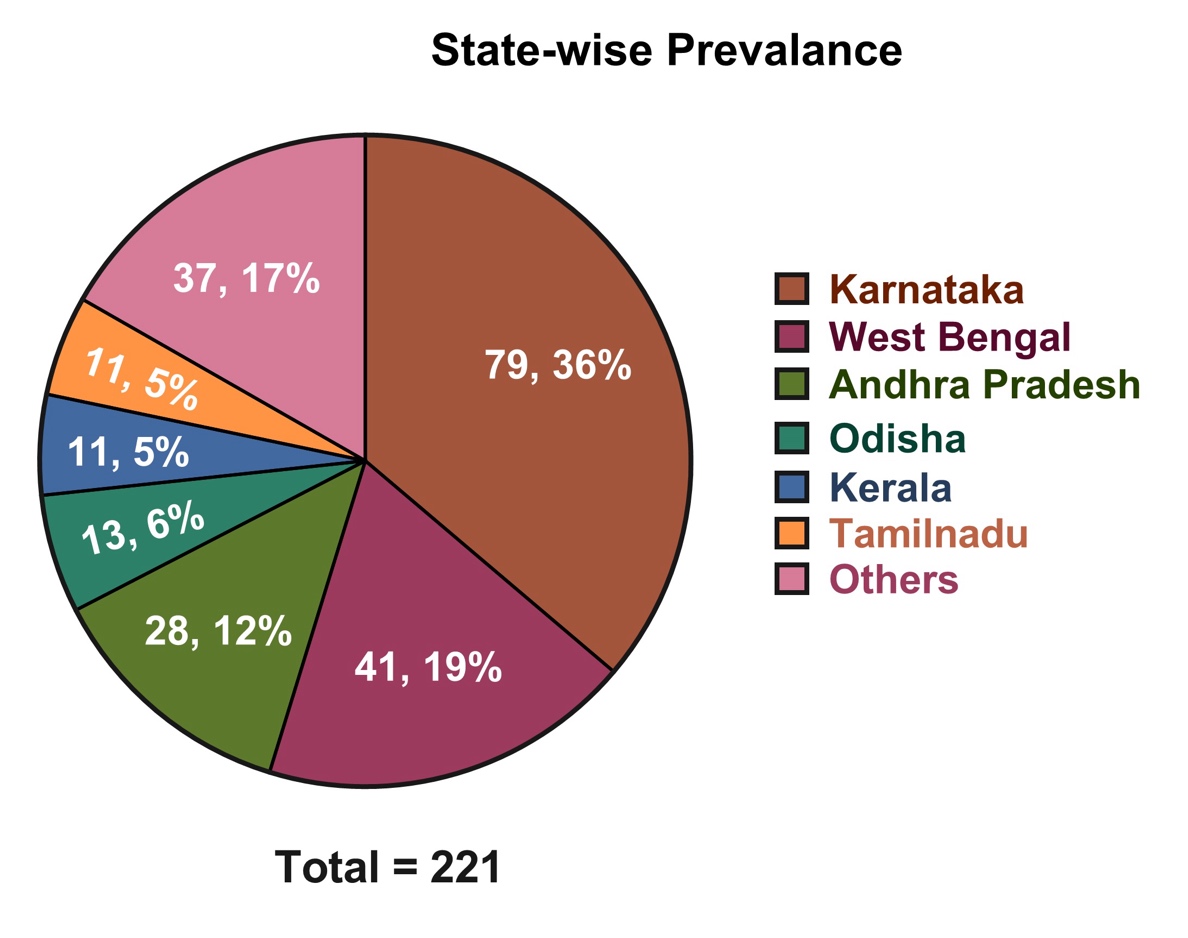

**Supplementary Figure 1:** State-wise distribution of the HD patients (n=220) (others: Bihar: 6 (2.7%), Uttar Pradesh: 4 (1.8%), Punjab: 4 (1.8%), Gujarat: 4 (1.8%), Assam: 4 (1.8%), Jharkhand: 3 (1.4%), Madhya Pradesh: 2 (1%), Maharashtra: 2 (1%), Tripura: 2 (1%), Delhi: 1 (0.5%), Five patients were from outside India, including 3 from Bangladesh and one each from Nepal and Iran

**Table 1: Frequency distribution of the longer and shorter allele CAG repeat lengths of HD patients (n=220)**

| **CAG repeat length** | **Count as longer allele** | **Count as shorter allele** | **Age at disease onset (years) mean ± SD (range)** |
| --- | --- | --- | --- |
| Up to 26 (normal) | 0 | 217 | .. |
| 27 – 35 (intermediate alleles) | 0 | 2 | .. |
| 36 – 39 (reduced penetrance) | 11 | 1 | 54.4 ± 10.8 (33 – 66) |
| 40 – 49 | 183 | 0 | 41.2 ± 10.2 (21 – 72) |
| 50 – 59 | 15 | 0 | 25.8 ± 4.8 (16 – 32) |
| 60 – 79 | 8 | 0 | 11.55 ± 3.1 (9 – 19) |
| 80 and above | 3 | 0 | 6.0 ± 2.6 (4 – 9) |

**Table 2: Frequency distribution of the age at disease onset of HD patients (n=220)**

| **Age at disease onset (years)** | **Number of patients** | **Longer allele CAG repeat mean ± SD (range)** |
| --- | --- | --- |
| < 10 | 5 | 84.4 ± 17.2 (68 -113) |
| 11 – 20 | 9 | 64.1 ± 7.6 (52 – 77) |
| 21 – 30 | 37 | 47.1 ± 4.5 (40 – 56) |
| 31 – 40 | 76 | 44.5 ± 2.4 (39 – 50) |
| 41 – 50 | 58 | 42.7 ± 1.8 (39 – 46) |
| 51 – 60 | 22 | 41.1 ± 1.4 (39 – 44) |
| 61 – 70 | 12 | 40.9 ± 2.2 (38 – 46) |
| 71 – 80 | 1 | 41.0 |

**Table 3: Details of parents and siblings of the HD cases without family history as informed by the patient/caregiver.**

| **ID** | **CAG count** | **AAO** | **Max age attained in years + relevant clinical info** | | **No. of siblings-status** |
| --- | --- | --- | --- | --- | --- |
|  |  |  | **Parent-1** | **Parent-2** |  |
| 79 | 23/43 | 41-45 | 41-45 | 76-80 | 3-unaffected |
| 64 | 18/42 | 51-55 | ~61-70 | ~61-70 | 5- unaffected |
| 29 | 15/40 | 51-55 | 76-80 | 51-55 | 2-unaffected |
| 50 | 16/44 | 41-45 | 96-100 | 71-75 | 4-unaffected |
| 55 | 18/43 | 36-40 | 61-65 | 71-75 | 3-unaffected |
| 46 | 19/44 | 36-40 | 56-60 | 56-60 | 3-unaffected (tested) |
| 45 | 17/41 | 36-40 | 61-65 | 71-75 | - |
| 22 | 22/41 | 46-50 | 91-95 | 96-100 | 5-unaffected |
| 80 | 17/41 | 56-60 | 81-85 | 71-75 | 3-unaffected  1-suicide |
| 44 | 17/40 | 46-50 | 66−70 | 66−70 | - |
| 34 | 17/45 | 40-45 | 51-55 | 61-65 | 5-unaffected |
| 74 | 17/41 | 51-55 | 66−70 | 66−70 | 7-unaffected  4*- late onset ~60* |
| 57 | 20/41 | 56-60 | 86−90 **(*mental illness*)** | 66−70 (Cardiac arrest) | - |
| 56 | 20/42 | 56-60 | 61-65 | 71-75 | 6-unaffected |
| 23 | 17/45 | 31-35 | 46−50 | 76-80 | 2-*affected*  *(but parents’ sibs unaffected)* |
| 66 | 18/42 | 46-50 | 56−60 | 56−60 | 2-unaffected |
| 66 | 18/44 | 36-40 | 56−60 | 56−60 | 5-unaffected |
| 24 | 17/41 | 51-55 | 66−70 | 46-50 | 6-unaffected |
| 30 | 17/42 | 61-65 | 66−70 | 71−75 | 3-unaffected |
| 55 | 18/43 | 66-50 | 66−70 | ~6−80 | 3-unaffected |
| 24 | 17/39 | 31-35 | 56−60* | 66−70 * | 4-unaffected |
| 75 | 17/40 | 61-65 | 61-65 | 61-65 | 2-unaffected |
| 24 | 20/40 | 36-40 | 71-75 | 81-85 | 6-unaffected |
| 27 | 16/50 | 26-30 | 66−70 * | 41-45 | 3-unaffected |
| 58 | 17/43 | 36-40 | 66−70 | 46-50 | 2-unaffected |
| 66 | 17/39 | 51-55 | 66−70 | 61-65  **(*mental illness*)** | 4-unaffected |
| 20 | 17/39 | 46-50 | 56−60 | 56−60 (***mental illness*)** | 8-unaffected |
| 30 | 17/48 | 21-25 | 66−70 * | 66−70 | 1-unaffected **(17/32)** |
| 50 | 17/41 | 56-60 | 76-80 | 56−60 **(*mental illness*)** | 3-unaffected  1-substance abuse |
| 26 | 17/43 | 36-40 | 66−70 * | 66−70* **(CAG 19/32)** | 1-unaffected |
| 32 | 22/45 | 31-35 | 56-60* | 66−70 * | 2-unaffected |
| 51 | **37/46** | 36-40 | 66−70 * | 31-35 (sudden death) | - |
| 20 | 18/44 | 51-55 | 76-80 | 76-80 | 2-unaffected,  1-*affected* |
| 21 | 17/44 | 36-40 | 66−70 * | 76-80* **(CAG 17/30)** | 3-unaffected |
| 41 | 18/41 | 56-60 | 56−60 | ~26-30 (unknown cause) | 3-unaffected  1-dementia(died) |
| 71 | 17/39 | 61-65 | 31-35 | 56−60 | 1-unaffected |
| 65 | 17/39 | 66-70 | 66−70 (***mental illness*)** | 66−70 | 8-unaffected  2-mental illness |

**Table 4: CAG-AAO trends in sibling pairs (n=23)**

| **Sl. No** | **Inheritance pattern** | **Elder sib CAG length** | **Younger sib CAG length** | **𝚫CAG*** | **Elder sib AAO** | **Younger sib AAO** | **𝚫AAO#** |
| --- | --- | --- | --- | --- | --- | --- | --- |
| 1 | Paternal | 47 | 43 | -4 | 31-35 | 36-40 | 1 |
| 2 | Paternal | 43 | 41 | -2 | 51-55 | 41-45 | -9 |
| 3 | Paternal | 43 | 44 | 1 | 46-50 | 41-45 | -2 |
| 4 | Paternal | 46 | 49 | 3 | 31-35 | 31-35 | 0 |
| 5 | Paternal | 46 | 45 | -1 | 31-35 | 36-40 | 7 |
| 6 | Paternal | 63 | 77 | 4 | 16-20 | 11-15 | -1 |
| 7 | Paternal | 42 | 49 | 7 | 61-65 | 31-35 | -28 |
| 8 | Paternal | 49 | 49 | 0 | 26-30 | 21-25 | -5 |
| 9 | Paternal | 38 | 43 | 5 | 61-65 | 41-45 | -19 |
| 10 | Paternal | 41 | 41 | 0 | 71-75 | 66-70 | -2 |
| 11 | Paternal | 43 | 41 | -2 | *36-40* | 31-35 | -3 |
| 12 | Paternal | 50 | 59 | 9 | 21-25 | 16-20 | -9 |
| 13 | Paternal | 85 | 80 | -5 | 1-5 | 6-10 | 4 |
| 14 | Paternal | 45 | 44 | -1 | 26-30 | 31-35 | 7 |
| 15 | Maternal | 42 | 43 | 1 | 51-55 | 51-55 | 3 |
| 16 | Maternal | 43 | 41 | -2 | 51-55 | 41-45 | -12 |
| 17 | Maternal | 42 | 41 | -1 | 51-55 | 41-45 | -9 |
| 18 | Maternal | 44 | 44 | 0 | 31-35 | 31-35 | 0 |
| 19 | Maternal | 47 | 45 | -2 | 26-30 | 31-35 | 3 |
| 20 | Maternal | 45 | 47 | 2 | *36-40* | 26-30 | -9 |
| 21 | Maternal | 43 | 46 | 3 | 46-50 | 31-35 | -15 |
| 22 | Unknown | 43 | 43 | 0 | 41-45 | 41-45 | -2 |
| 23 | Paternal | 45 | 43 | -2 | 36-40 | NA | NA |
|  | Mean ± SD (range) | 46.7 ± 9.58 (38 - 85) | 47.73 ± 10.5 (41 – 80) |  | 40.18 ± 16.1 (5 – 72) | 35.6 ± 13.4 (9 – 70) |  |
| *Delta CAG = Younger sib CAG length – Elder sib CAG length  #Delta AAO = Younger sib AAO (years) – Elder sib AAO (years); AAO = Age at disease onset. Italicized are not included in analysis due to pre-manifest individuals in the pair | | | | | | | |

**Table 5: CAG-AAO trends in parent-offspring pairs (n=13)**

| **Sl. No** | **Inheritance pattern** | **Parent CAG length** | **Child CAG length** | **𝚫CAG*** | **Parent AAO** | **Offspring AAO** | **𝚫AAO #** |
| --- | --- | --- | --- | --- | --- | --- | --- |
| 1 | Paternal | 43 | 43 | 0 | 51-55 | 31-35 | -18 |
| ~~2~~ | Paternal | 44 | 64 | 20 | 41-45 | 16-20 | -25 |
| 3 | Paternal | 48 | 62 | 14 | 31-35 | 11-15 | -18 |
| 4 | Paternal | 42 | 45 | 3 | 61-65 | 21-25 | -39 |
| 5 | Paternal | 49 | 77 | 28 | 31-35 | 11-15 | -22 |
| 6 | Paternal | 49 | 63 | 24 | 31-35 | 16-20 | -15 |
| 7 | Paternal | 41 | 51 | 10 | 71-75 | 26-30 | -42 |
| 8 | Maternal | 50 | 69 | 19 | 26-30 | 11-15 | -16 |
| 9 | Maternal | 43 | 41 | -2 | 51-55 | 26-30 | -27 |
| 10 | Maternal | 44 | 45 | 1 | 41-45 | 31-35 | -7 |
| *11* | *Paternal* | *43* | *113* | *70* | *NA* | *1-5* | *NA* |
| *12* | *Maternal* | *42* | *41* | *-1* | 61-65 | *NA* | *NA* |
| *13* | *Maternal* | *42* | *42* | *0* | *41-45* | *NA* | *NA* |
|  | Mean ± SD (range) | 45.1 ± 3.1 (41 - 50) | 61.1 ± 20.4 (41 – 113) |  | 45.6 ± 14.5 (30 – 72) | 21.0 ± 8.7 (11 – 35) |  |
| *Delta CAG = Offspring CAG length – Parent CAG length  #Delta AAO = Offspring AAO (years) – Parent AAO (years); AAO = age at disease onset  Italicized are not included in analysis due to pre-manifest individuals in the pair | | | | | | | |

**Table 6: Intermediate allele (IA) carriers**

| **s no** | **Intermediate allele carrier** | **Genotype** | |
| --- | --- | --- | --- |
|  |  | Longer allele | shorter allele |
| 1 | Parent of a de novo expansion carrier | 32 | 19 |
| 2 | sibling of a de novo expansion carrier | 32 | 17 |
| 3 | HD phenocopy | 31 | 9 |
| 4 | Parent of a de novo expansion carrier | 30 | 17 |
| 5 | HD phenocopy | 30 | 16 |
| 6 | HD phenocopy | 30 | 16 |
| 7 | HD patient | 43 | 28 |
| 8 | HD patient | 47 | 27 |
